# Differential Impacts of the COVID-19 Pandemic on Sociodemographic Groups in England: A Mathematical Model Framework

**DOI:** 10.1101/2024.11.09.24317003

**Authors:** Gbeminiyi J. Oyedele, Ivo Vlaev, Michael J. Tildesley

## Abstract

The Coronavirus disease-2019 (COVID-19) pandemic has had a significant impact on the world, redefining how we work, respond to public health emergencies and control efforts, and sparking increased research efforts. In this study, we have developed a deterministic, ordinary differential equation multi-risk structured model of the disease outcomes, with a focus on the total number of infections, reported cases, hospitalised individuals, and deaths in the population. The model takes into account sociodemographic risk-structure and age structured dynamics, as well as time-sensitive nonpharmaceutical interventions (lockdowns) to help observe the disease trajectory following the implementation of control measures. The primary focus of this study is to demonstrate the impact of different patterns of social mixing within and between deprivation deciles in England, to understand disparities in disease outcomes. Our analysis reveals that the diagonal kind of mixing, similar to “within-group homogenous” type of mixing assumption, results in a higher number of disease outcome compared to other types of mixing assumptions. We also explore the effectiveness of movement restriction (the first national lockdown) in controlling the spread of the virus in each social group, in order to understand how to target interventions in the future. Our analysis confirms significant disparities in infection outcomes between sociodemographic groups in England.

**Author summary:** The global impact of the coronavirus pandemic 2019 was evident, but different sociodemographic groups experienced disproportionate disease outcomes. In this paper, we present results from a mathematical model that simulates COVID-19 outcomes across diverse sociodemographic groups in England. Our work uses a mathematical framework that combines age and deprivation decile, to examine the disproportionate outcome in the number of infection, hospitalisation, and mortality based on social mixing patterns. Our work demonstrated the elevated risk for more deprived groups, where social and occupational factors increase contact rates, therefore intensifying disease spread. By distinguishing disease dynamics among deprivation deciles, this model offers insights for policymakers to design more equitable health strategies. This approach emphasis the need for policies that address the vulnerabilities of specific social groups to mitigate the effects of pandemics.

## Introduction

In late 2019, the world experienced a new virus – Severe Acute Respiratory Syndrome Coronavirus–2 (SARS-CoV-2) – of pandemic magnitude that redefined how people worked and responded to public health emergencies. During the global spread of SARS-CoV-2, non-pharmaceutical interventions (NPI) were widely implemented as one of the measures to relax virus transmission. Measures such as stay-at-home orders, social distancing, hand washing, school closure, closures of social and religious centres, and mask wearing were some of the first NPIs measures implemented, after which other pharmaceutical interventions such as vaccination and management drugs, among others were adopted to slow down COVID-19 transmission globally [1]. Research such as [2] has demonstrated that the differences in the incidence and severity of COVID-19 between countries can be attributed to the population’s reaction to the NPIs that were implemented.

Although the virus had a global impact, differential outcomes were reported across sociodemographic groups [3–5] highlighting that the COVID-19 pandemic affected different parts of the population disproportionately. A notable instance is the disproportionately high rate of COVID-19 infection, hospitalisation, and mortality among racial and ethnic minorities [4]. Ethnic minorities, despite making up only 14% of the population in the UK, account for 34% of those critically ill [6]. This differential outcome could be attributed to disparities in healthcare access and socioeconomic opportunities between the majority and minority ethnic groups. Therefore, it is crucial to examine the differential impacts of the pandemic on various sociodemographic groups, to enable policymakers to identify and address these disparities effectively. We seek to examine how social mixing patterns influences the varied impacts of the COVID-19 pandemic across different sociodemographic groups in England, United Kingdom (UK). In this study, sociodemographic groups are defined using the 2019 Index of Multiple Deprivation (IMD) [7], which serves as the recognized metric for assessing relative deprivation in England. Research has suggested that social and health inequalities are associated with level of deprivation [8], and [9] have shown that deprivation is a major risk factor in the mortality rate of COVID-19. Such differences in COVID-19 disease outcomes based on deprivation levels may stem from economic, social, and health inequalities.

For instance, the population of people in the most deprived groups may face challenges working remotely, rely more on public transport, and may experience increased contact in workplace settings compared to those in the least deprived groups, thus increasing their risk of contracting a disease such as COVID-19. To reduce these disparities, it is necessary to identify factors such as age, jobs, income, etc. that increase risk within each deprived group. As a result, infectious disease models have become increasingly important for policymakers in providing solutions to problems related to disease outbreaks and epidemiological issues, especially during endemic or epidemic health crises [10]. Mathematical models, in particular, have helped in providing epidemiological insights and have been utilised to estimate epidemic size, incidence, the size of the infection peak, and the impact of intervention policies.

Some mathematical models have been used to highlight the significance of age in disease spread, both in terms of case incidence and disease severity during the pandemic [11–13]. It has been reported that there are disparities in the severity and mortality of the virus based on age [11]. In addition, susceptibility, transmissibility, hospitalisation rates and death rates have been shown to be age-dependent [12, 14, 15]. The study by [12] revealed that susceptibility of individuals under 20 years of age to coronavirus disease is approximately half that of adults aged over 20 years. Furthermore, it was found that clinical symptoms (requiring hospital care) occurred in 21% of infections in 10-19 year olds, compared to 69% of infections in people aged over 70 years. The study also suggests that interventions targeted at children may have a limited effect reducing SARS-CoV-2 transmission, particularly if the subclinical transmissibility of the infection is low [12]. The COVID-19 pandemic’s impact varied across different communities and individuals. Densely populated areas and jobs requiring frequent contact (healthcare and service industry) elevated infection risk, particularly for lower socioeconomic status (SES) communities. Manna et al. (2023) [16] introduced a model creating synthetic generalized contact matrices that incorporate age, SES such as income, employment, and gender for epidemic modeling. A study by Manna et al. (2023) [17] in Hungary demonstrated that employment status and education significantly affected contact rates and vaccination acceptance, highlighting social determinants’ role in epidemic-related behaviours. Hale et al. (2022) [18] used the 2019 English Index of Multiple Deprivation (IMD) to segregate the population based on deprivation levels, showing that variations in social mixing, testing/reporting motivation, and precautionary measures (e.g., mask-wearing, vaccination) contributed to differences in daily contact rates between the most and least deprived groups.

In this paper we expanded upon the concepts introduced by [18] and [16] to create a deterministic mathematical modelling framework capable of analysing disease dynamics across different age and deprivation decile. The objective of this study is to develop a deterministic mathematical model framework that would help study the effect of social mixing within and between deprivation deciles in England. This will provide insight into the mechanisms resulting in disproportionate disease outcomes between different sociodemographic groups during the COVID-19 pandemic.

## Materials and methods

### Model Description

In this study, we use the deterministic model of ordinary differential equations in our previous work [19]. The model incorporates age and sociodemographic contact mixing patterns and is parameterised using existing literature to understand the dynamics of infection, hospitalisation, and mortality of COVID-19 in England. The social and age groups were subdivided into an extended compartmental SEIR-type mathematical model. Sociodemographic groups were defined as deprivation deciles based on the Index of Multiple Deprivation (IMD), a measure extrapolated from seven domains of deprivation: income, employment, education, health, crime, barriers to housing and services, and living environment [7]—and used to define social groups at the lower-layer Super Output Area (LSOA) level or neighbourhood. Four or five groups of output areas (OAs), each comprising 40 to 250 households, make up the LSOAs [20]. A typical LSOA will include between 400*and*1, 200 households, with a resident population of between 1, 000 and 3, 000 [20].

In England, the indices of multiple deprivation (IMD) are divided into (32, 844) distinct categories, with rankings ranging from the most deprived group (1) to the least deprived group (32, 844). This total was divided into ten equal segments to form deprivation deciles. The mathematical model framework proposed in this study divides each sociodemographic group (deprivation deciles) into 21 age classes, segregated by 5-year age band: 0-4, 5-9,…, 90+. For each “social-age group”, the total population 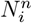 at time *t* is given as:

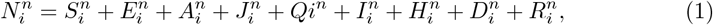

Susceptible individuals 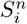 include those who have never had the disease, as well as those whose immunity has waned post-recovery, necessitating reinfection modelling. Exposed individuals 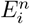 have been in contact with an infected individual but are not yet infectious. Some exposed individuals are tested for SARS-CoV-2 virus, while non-tested individuals proceed to either symptomatic, asymptomatic, or symptomatic reported compartments. Asymptomatic individuals 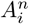 can spread the disease without showing symptoms. Symptomatic non-reported individuals, 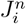, show symptoms, can infect others, but are untested, contributing to case underreporting and may require hospitalisation if severe. Symptomatic tested individuals 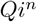 wait 3 days for results, transmitting at a reduced rate; if negative, they recover post-infectious period.

Reported cases 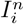 have tested positive, can spread infection, and might possibly need hospital care. Hospitalized individuals are 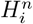 Dead individuals are 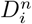. Recovered individuals 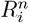 can become susceptible again as their immunity wanes.

The model assumes that susceptible individuals contact infectious individuals at rate *β*_*n,i*_(*t*), a function of social mixing, age mixing, susceptibility and infectivity by age and relative transmission rate. Upon contact with infectious individuals they transition into the exposed compartment. Exposed individuals progress to the asymptomatic or unreported symptomatic compartment if untested for COVID-19 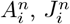.Others take a test (PCR or lateral flow) and move to compartment 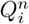.In compartment 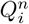, individuals are tested at rate 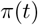 and wait for an average period 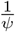 *days* for results. In this study, we make the assumption that the testing capacity grows over time, and is very low at the outbreak’s start, aligning with [18]. Parameter *π*(*t*), which is the testing capacity is defined by the Generalised Richards Model (GRM) proposed by [21], is defined in equation (2).

The cumulative number of tests by time is given as:

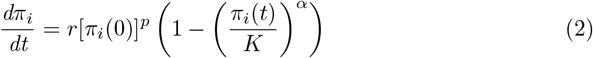

where *r* is the growth rate, *K* is the carrying capacity, *p* is the growth profile (that is, if *p* = *α* = 1 we would have the classical logistic growth model), *α* is the deviation away from the S-shape of the logistic curve, and *π*_*i*_(0) is the initial number of tests. Here, we assume test rate is uniform and the same in the whole population. We estimated the parameters *r, p, K, & α* using the Non-linear least square method, by fitting to the UK COVID-19 cumulative number of tests data from January, 03 - September, 03 2020 of the publicly available data [22]. This study considered the period when the wild-type of SARS-CoV2 was dominant in the UK, and the period of the first lockdown.

The model equations are defined as follows:

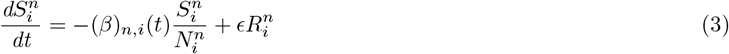

[eq

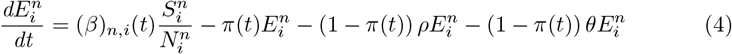

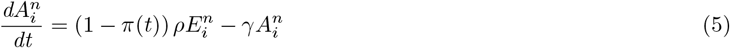

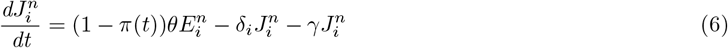

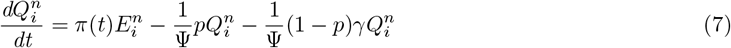

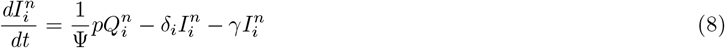

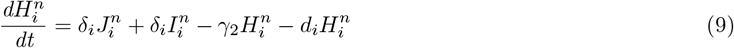

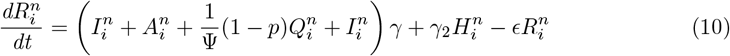

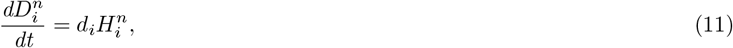

and *θ* = 1 –*ρ*.

The force of infection is defined as the effective contact between and within deprivation deciles (while maintaining constant age mixing) that produces an infection. Previous studies have shown the differential impact of age on the dynamics of Coronavirus 2019 (COVID-19) [13, 23, 24]. Mathematically, we define the force of infection as:

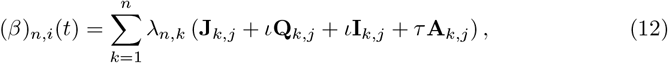

and *λ* = (*λ*_*n,m*_) as a block matrix, where *n, m* = 1, 2, · · ·, 10. Each element (*λ*_*n,m*_) of this block matrix is a 21 × 21 age mixing matrix. Therefore we express *λ*_*n,m*_ as:

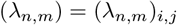

where *i, j* = 1, 2, …, 21.

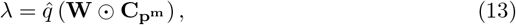

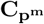 is a vector containing the age-mixing for each deprivation decile, **W** is the social mixing matrix, and 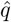 is the relative transmission rate. Each **C**_**i**_ (*for i* = 1, · · ·, *p*^*m*^) is an age mixing matrix, and 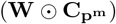 is an elementwise multiplication of the form:

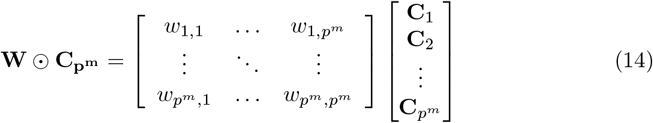

Due to unavailability of data related to the age mixing matrix by the deprivation decile, we assumed 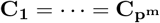. Then

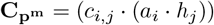

From equation 13, *C*_*k*_ is defined as the age mixing in each deprivation decile (which we assume to be equal across decile), and the element in each *C*_*k*_ is *c*_*i,j*_ which is the average number of contacts per day from POLYMOD data [25], *a*_*i*_ the age-related differential infectivity, and *h*_*i*_ the age-related differential susceptibility. The POLYMOD study is a large-scale quantitative study to understand contact patterns for infectious diseases transmitted by respiration or close contact [25]. This study used scaled POLYMOD data for England segmented into 21 age groups (5-year age bands) as used in the work of [13]. The contacts were divided into an average number of contacts per day at home, school, work, and other places. The total number of contacts is defined as the sum of contacts at home, school, work and other places:

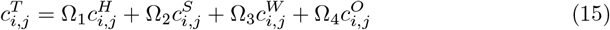

where Ω_1−4_ in equation 15 are the scaling factor for the age mixing during the lockdown period.

The mixing pattern by deprivation decile (sociodemographic groups) **W** is defined by the equation proposed by [26]. Each element *w*_*n,m*_ in **W** is defined as the average number of contacts made within and between the deprivation deciles given as:

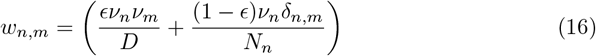

where (δ_*n,m*_ : *m* = *m is* 1, *& n* ≠ *m is* 0), *ϵ* ∈ [0, 1], and ν_*n*_ is the average number of contacts by each sociodemographic group, *N*_*n*_ is the total population by decile in England. *D* = Σ_*n*=1_ν_*n*_*N*_*n*_ is the total number of contacts per unit time made by all people in the population. The element of each social mixing matrix **W** is generated by normalising the equation (16) so that the sum of the matrix **W** is equal to 1. We normalised this by dividing **W** by 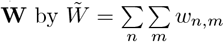,which is the sum of all contacts generated by the sociodemographic groups.

To our knowledge, no contact survey on the social mixing matrix has been identified by deciles of deprivation. Few studies such as [18, 26, 27] that studied mixing between different demographic groups had to make some assumptions about the mixing pattern of the population they were studying. We made a simplistic assumption that the average number of contacts in each sociodemographic group decreases linearly from 10 to 1, with 10 representing the daily average number of interactions for the most deprived group and 1 for the least deprived. We then adjusted the value of *ϵ* to examine various mixing patterns. Our aim is to investigate the impact of different mixing patterns on disease outcomes between sociodemographic groups. We study the diagonal (people mix only within their groups and nowhere else), preferred (people are more likely to interact with those in their own group) and proportionate (people mix randomly with the entire population) types of mixing using the defined mixing assumptions.

### Lockdown Implementation

In this study, we made the simplified assumption that lockdown reduced socio-contact mixing by 50% within the decile of deprivation. This is to capture the impact of lockdown on employment-related contact across the deprivation decile; it was assumed that essential workers, who remained physically present at work, may contribute to ongoing transmission. Deprivation is known to be correlated with age [18], allowing us to focus more on the lockdown strategy within the age mixing matrix. The implementation of the restriction by age is presented in table 1.

**Table 1.** Lockdown implementation by age groups.

| Restriction | Description | value (equation 15) |
| --- | --- | --- |
| Total Lockdown | Closing non-essential places like pubs and supermarkets boosts home mixing and lowers workplace mixing. Home mixing adjusts for higher household transmission during lockdown by a factor $\Omega_1$, while $\Omega_3$ indicates minimal workplace mixing. | $c_{i,j}^T = c_{i,j}^H \Omega_1 + c_{i,j}^W \Omega_3$ |
| Easing | As more non-essential places and workplaces open, workplace interaction increases, scaling up the parameter $\Omega_3$, and some minimal interaction in other places $\Omega_2$. | $c_{i,j}^T = c_{i,j}^H \Omega_1 + c_{i,j}^W \Omega_3 + c_{i,j}^O \Omega_4$ |
| Relaxed Restriction | Everything that happened during easing of lockdown with the addition of minimised mixing at other places by the scaling parameter $\Omega_4$. The removal of restrictions will mean that all locations are accessible again, however, the rate of mixing has not yet returned to what it was prior to the lockdown. | $c_{i,j}^T = c_{i,j}^H \Omega_1 + c_{i,j}^W \Omega_3 + c_{i,j}^S \Omega_2 + c_{i,j}^O \Omega_4$ |

## Results

In this study, parameters were selected from the existing literature (see table 2) and simulated for 270 days with the assumption of observing the period when the wild-type SAR-CoV2 was still dominant and the period when the first lockdown and easing was implemented. We assume that a reported infected individual entered the system *I*(0) = 1 in the most deprived group and age group 55 − 59. This assumption was maintained for all variations of *ϵ* except for when *ϵ* = 0, when we assumed *I*(0) = 1 for all deciles of deprivation but the same age group. The focus was to determine the total number of infections, reported cases, hospitalised individuals, and deaths. Thereafter, the infection trajectory, hospitalisation, and death by the decile of deprivation were presented. The findings illustrate the influence of integrating different mixing scenarios both within and across deprivation deciles. In this analysis, the cumulative number of infections at any specific time (*t*) was determined by summing the individuals in the reported and unreported symptomatic, asymptomatic, and hospitalised categories.

**Table 2.** Model parameter values and references.

| Parameter | Description | Value & Source |
| --- | --- | --- |
| $c_{i,j}$ | contact matrix by age | [ [25]] |
| $\iota$ | The modification factor by which confirmed and reported individuals can transmit infection. | 0.27 [ [13]] |
| $\rho$ | Proportion of population that are asymptomatic | 0.6 |
| $\theta$ | Proportion of population that are symptomatic | 0.4 [ [28]] |
| $p$ | Sensitivity of test, which we used as the probability of testing positive to test | 0.7826 [ [28]] |
| $\Psi$ | The average number of days for test result to be reported | 1/3 *days*<sup>-1</sup> (assumed) |
| $q$ | Relative infection rate | 0.9 (assumed) |
| $\gamma$ | recovery rate | 1/14 *days*<sup>-1</sup> [ [28]] |
| $\delta_i$ | The rate of symptomatic or positive tested individuals become hospitalised, varies by age. | [ [13]] |
| $d_i$ | The rate at which hospitalised individual become fatally sick. This varies by age | [ [13]] |
| $\epsilon$ | probability of wanning immunity | 2/100 *days*<sup>-1</sup> [ [29]] |
| $a_i$ | Infectivity by age | [ [13]] |
| $h_i$ | susceptibility by age | [ [13]] |

Reported cases were defined as those who had tested positive for the SARS-CoV2 virus after taking a test and were considered confirmed cases in the population. In the figures presented, the red vertical line corresponds to the lockdown period on March 23, 2020, and the green vertical line corresponds to the lockdown easing period from June 15, 2020.

### Diagonal Mixing Scenario

Diagonal mixing occurs in our model when *ϵ* = 0. In this scenario, individuals exclusively engage with others within their own group, without any interaction with individuals from different social groups. This results in a “within-group homogeneous” mixing pattern, where each group operates independently of the others, indicating that the individuals within each decile mix evenly and possess similar traits. As a result, Equation 12 can be expressed as:

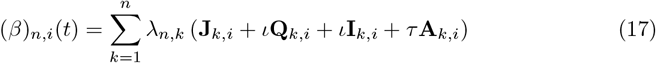

and *λ* remains as equation 13, however the hazard rate is now defined for each deprivation decile. Equation 17 demonstrates that the growth of infection occurs independently within each group. This is because there is no interaction between the groups; only within-group dynamics is observed. Since there is no interaction between deciles, we initialised our model by introducing an infection into each decile and followed our initial assumption that the most deprived groups tend to mix more frequently than people in the least deprived groups. This assumption about social mixing behaviour is reasonable considering that people in the most deprived groups are likely to have more interactions due to factors such as the reliance on public transportation, the size of the household and the type of employment [18].

Consequently, to observe an infection trajectory, there must be at least one infectious individual present in each sociodemographic group.

This work is built on the rigorous mathematical analysis of the model presented in the work of [19]. The findings indicate that the *R*_0_ values were significantly elevated in the groups with increased social interactions in contrast to the groups with limited interactions, supporting the previous hypothesis that the *R*_0_ value is influenced by the structure of contacts and the behavioural dynamics of the population [30].This suggests that the group exhibiting a notably high level of interaction creates sufficient mixing to produce secondary infections, as depicted in Figure 1 and 2. Since mixing is only observed within deciles (no mixing between deciles), the value of *R*_0_ must be greater than 1 in each decile for disease to spread (or generate secondary infections); otherwise, the infection will die out because on average an infectious individual cannot infect at least one other person when *R*_0_ *<* 1. Consequently, disease transmission is expected to be more prevalent in high-activity groups compared to those in the least deprived areas.

**Fig 1.**
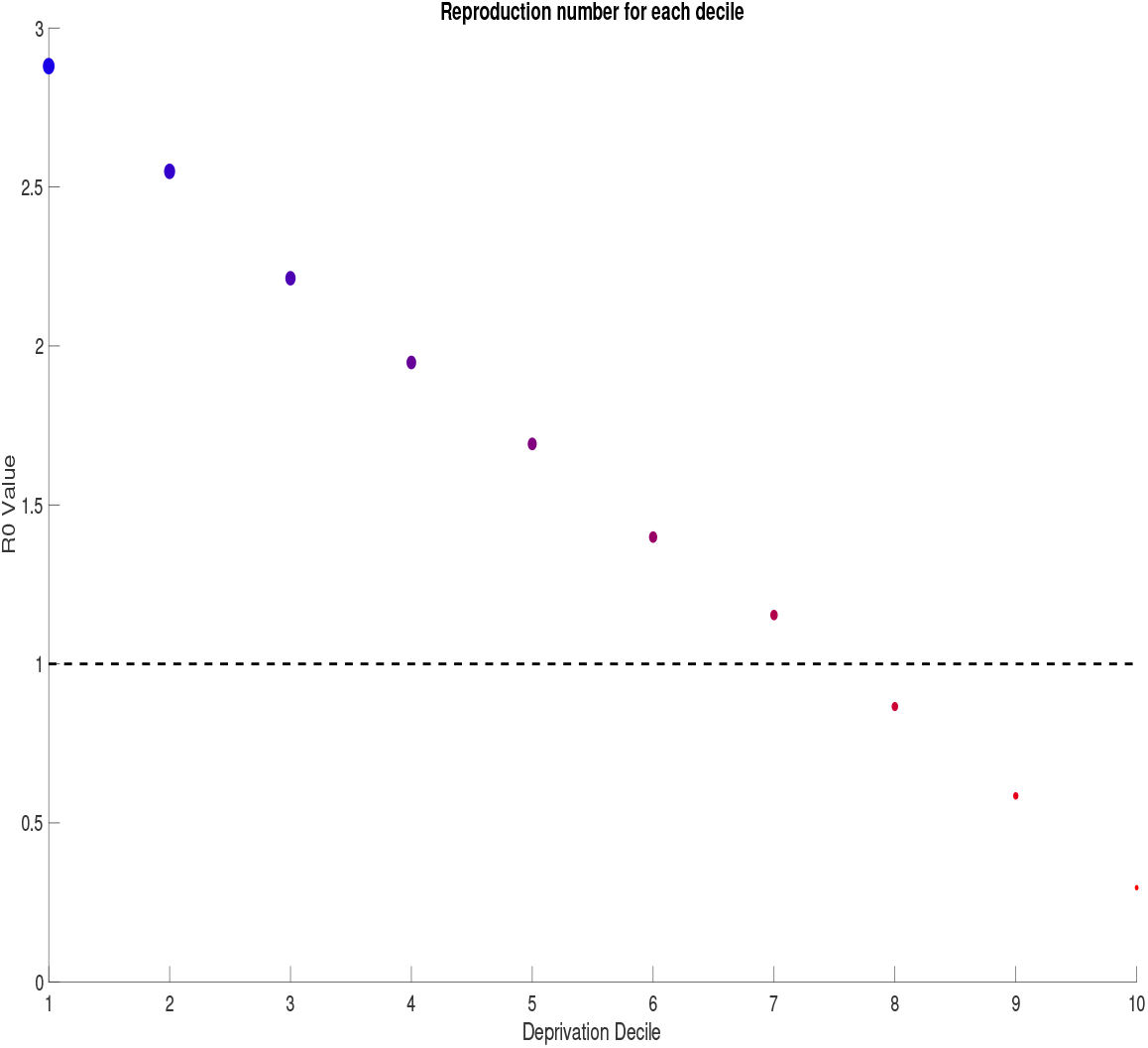
Basic reproduction number (R0) for each deprivation decile, demonstrating the variation in disease transmission potential across social groups. Higher R0 values observed in more deprived groups is due to increased social mixing and contact rates. The figure highlights the significant disparity in transmission dynamics influenced by sociodemographic factors.

**Fig 2.**
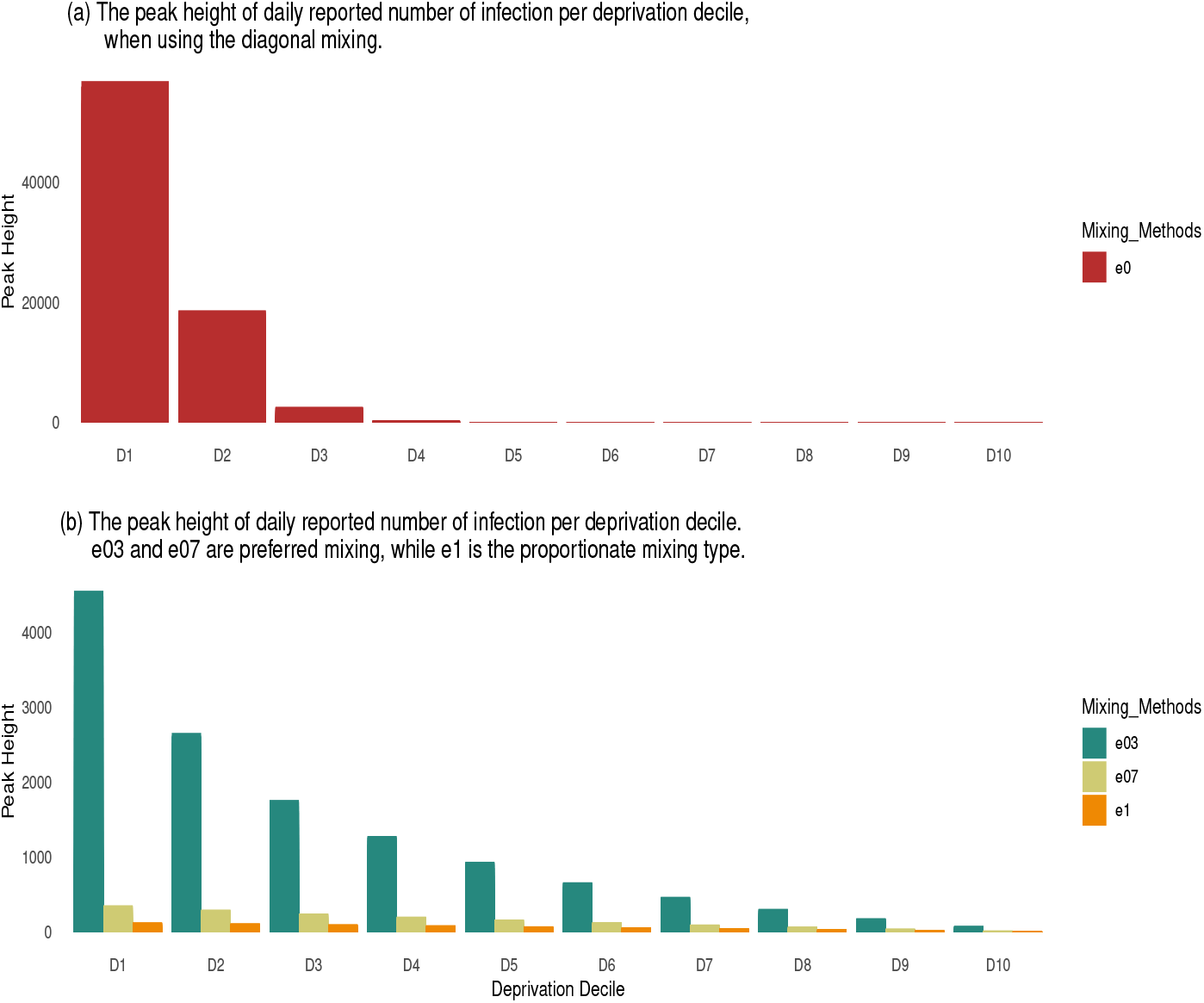
Comparison of peak height of infection per deprivation decile. Top panel: peak height of infection by deprivation decile for the diagonal mixing assumption (*ϵ* = 0). Bottom panel: Combined the preferred (*ϵ* = 0.3 *and ϵ* = 0.7) and proportionate (*ϵ* = 1) mixing assumptions for the peak infection height in each deprivation decile. This figure illustrates how different social mixing patterns affect the infection burden across deprivation levels, with more mixing within groups driving higher peak infections in the most deprived deciles.

### Preferred Mixing Assumption

This mixing scenario is suitable for systems where individuals in the same group are more likely to interact than to engage in “random” mixing between different groups. The preferred type of mixing is designed to demonstrate interaction heterogeneity in sociodemographic groups, by assuming that there is more likelihood of contacting individuals in your social group as compared to randomly meeting people outside your social group. In this study, we assumed two values for *ϵ*: (i) *ϵ* = 0.3 (stronger mixing or interaction within groups and significantly minimal mixing between groups) and (ii) *ϵ* = 0.7 (strong mixing within groups and significant mixing between the upper most deprived groups).

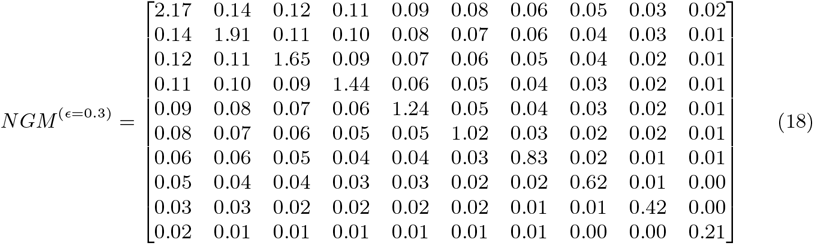

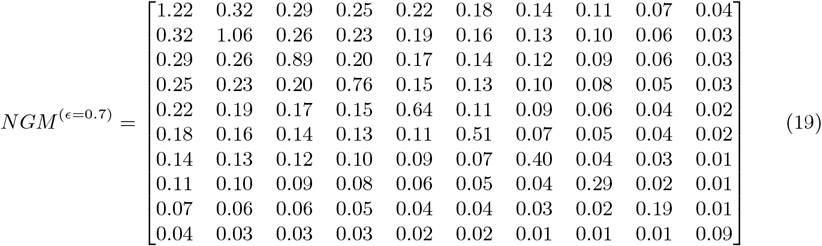

The values of *ϵ* varied between 0 and 1 as shown in the next-generation matrix equation (18) and (19) denotes the intensity of mixing along the main diagonal. The preferred type of mixing assumes frequent interactions within similar groups rather than between different groups. This kind of interaction can be applied to different types of social interaction in human population. For example, we would expect someone in an executive position who is most likely to be in the least deprived group to interact frequently with people in the least deprived group and less likely to interact with people in the most deprived group. Hence, this kind of setup captures the strong mixing within the main diagonal and linearly decreasing interactions outside the main diagonal (meaning that the farther you are from a group, the less likely it is for you to interact with that group).

By comparing the disease progression derived from equations (18) and (19), it is evident that the greater interaction along the main diagonal in (18) led to a higher incidence of infections, as depicted in Figure (3), in contrast to Figure (4) which shows a lower infection outcome. This difference is particularly noticeable in the markedly elevated number of secondary infections that can arise within the most deprived when *ϵ* = 0.3 (indicating more intense mixing along the main diagonal) compared to the scenario when *ϵ* = 0.7. Unlike the situation with diagonal mixing, where a higher infection rate was observed due to interactions solely within the same group, the infection diminishes more rapidly even after the relaxation of containment measures (lockdown).

### Proportionate Mixing Assumption

At *ϵ* = 1 in equation (16) we will observe proportionate mixing, where the mixing within and between sociodemographic groups are equally likely. This means that people could simply interact with anyone without taking into account their social domain or status. However, even with this “equal likelihood” in mixing, social groups still tend to interact with groups closer to themselves compared to those far from them on the social spectrum. This form of mixing can facilitate interactions that resemble “spill-over” between groups without a specific focus on connecting with more individuals from one’s own demographic group. Also, when *ϵ* = 1, equation (16) only takes into account the quadratic part, so there is no forcing on the main diagonal. This implies that the values along the main diagonal are squared by the average number of contacts in each group and multiplied by the average number of contacts with other groups off the main diagonal. Consequently, contact decreases as it moves away from the most deprived group towards the least deprived group, but that does not necessarily mean that the mixing along the diagonal is more than the mixing off the diagonal.

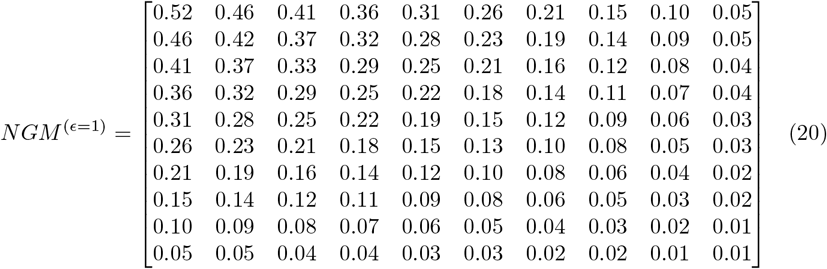

Analysis ofthe next generation matrix in equation (20) indicates that an infectious individual in the most deprived group would effectively contact 2.83 individuals on average (compared with effective contact of 0.28 in least deprived group), of whom 0.52 are people of the same decile group, 0.46 are the next deprived group (decile 2) and so on. However, as individuals in the most deprived groups mixed with equal likelihood with individuals in other groups in “random-like” interactions, the spread of disease dynamics could become more consistent across all groups.

## Discussion

In this study, we developed a modelling framework that combined mixing pattern of age and deprivation decile of the population of England to understand COVID-19 outcomes such as infection, hospitalisation, and deaths. Our findings reveal that diagonal mixing results in a higher number of disease outcomes compared to other types of mixing assumptions. All mixing assumptions that are presented agree that the most deprived group has a disproportionately higher rate of infection outcome (incidence, hospitalisation, and mortality) during the outbreak. This suggests that both socioeconomic status and social interaction patterns are critical factors influencing disease transmission and severity.

Previous research [16–18, 31–34] has emphasised the potential influence of sociodemographic groups such as age, education, and income on the outcomes of infectious diseases. In recent times, only a few of these studies [16–18] have actually coupled age and other sociodemographic groups into a mathematical model to understand the dynamic of infectious diseases, and one [18] used the deprivation decile. Although our model findings align with the mathematical model results of [18], a shift known as “deprivation switching” was observed in their work. This shift involved the least deprived group exhibiting higher infection outcome compared to the most deprived group during the spring 2021, before switching back to the most deprived group exhibiting a higher infection outcome similar to the earlier stage of the pandemic. This phenomena could be explained as either change in reporting behaviour across deprivation group or increased variation in social mixing across deprivation decile [18].

During the spring of 2021, there was a noticeable shift where the least deprived group showed a higher rate of infection outcomes compared to the most deprived group. Subsequently, the trend reverted to the earlier pattern, with the most deprived group having higher infection rates. This occurrence may be attributed to either change in reporting behaviour among different deprivation groups or enhanced variation in social interactions across deprivation decile [18]. This occurrence underscored the importance of considering time periods, social interaction, and behavioural change when constructing mathematical models.

In our study, the disease outcomes in the diagonal mixing scenario are higher compared with other mixing scenarios. The preferred mixing scenario, where similar groups tend to engage with each other frequently, the high-activity group, which represents the most disadvantaged groups in our case, will predominantly interact within their own group rather than with other groups. This leads to notably elevated rates of disease outcomes within those groups, as illustrated in Figures 3 and 4. Thus, promoting the focus of control efforts on the most deprived groups of the population is crucial. This is because when the most deprived groups achieve herd immunity and the value of *R*_0_ in that group falls below 1, the capacity to spread secondary infections decreases. As a result, the probability of infection spill-over to other deprivation deciles is ultimately reduced, benefitting the overall population.

**Fig 3.**
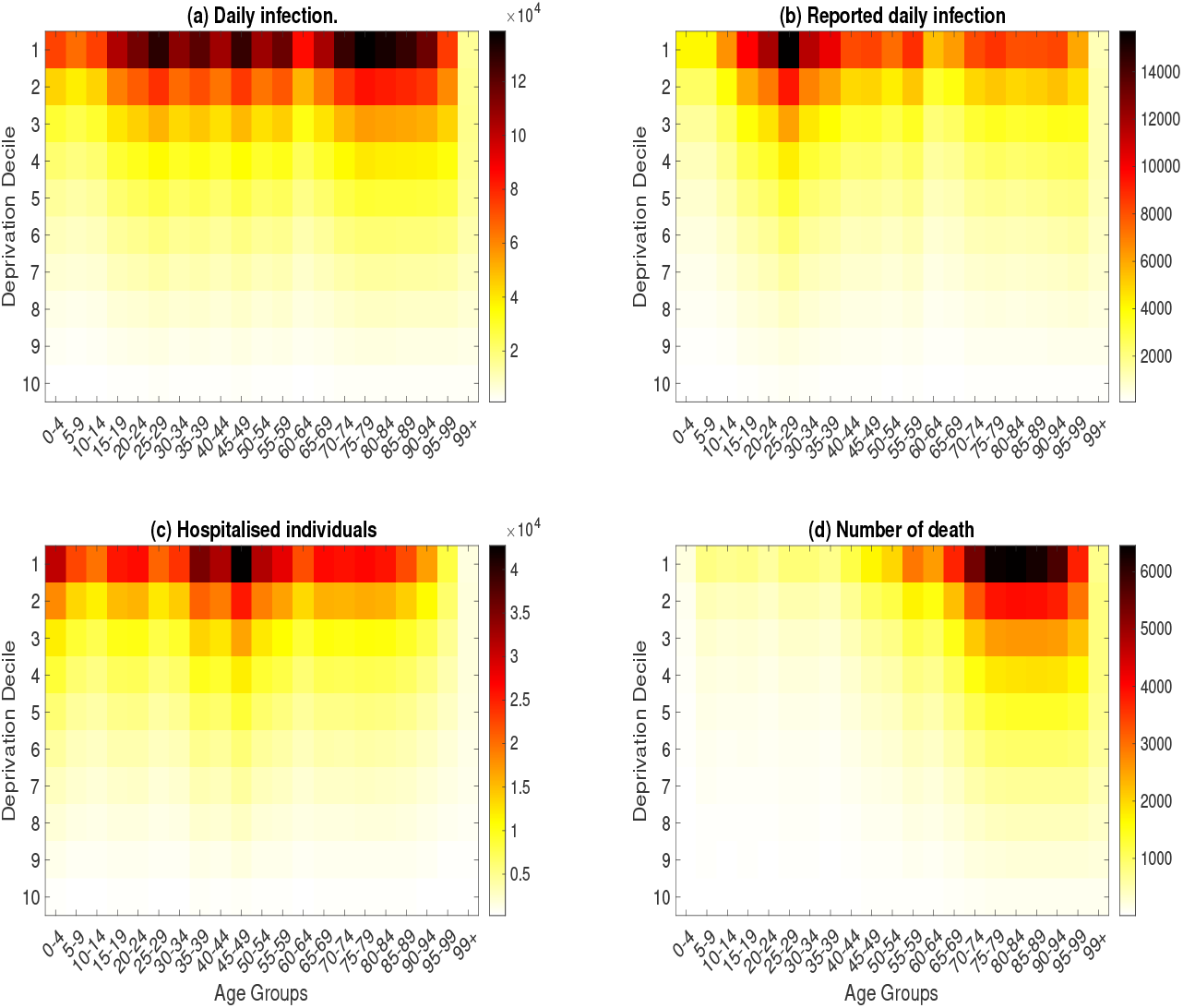
Top left panel: sum of the daily number of infection aggregated by age and deprivation decile. **Top right panel:** sum of the daily number of reported cases aggregated by age and deprivation decile. **Bottom right panel:** The sum of hospitalised individuals by age and deprivation decile. **Bottom left panel** The sum of daily death by age and deprivation decile. This figure highlights the unequal burden of COVID-19 outcomes across age and deprivation groups at *ϵ* = 0.3. This shows a greater concentration of severe outcomes in older age groups and most deprivation deciles. The disparities are more pronounced in most deprived deciles, reflecting the combined impact of age and socioeconomic factors on disease severity.

**Fig 4.**
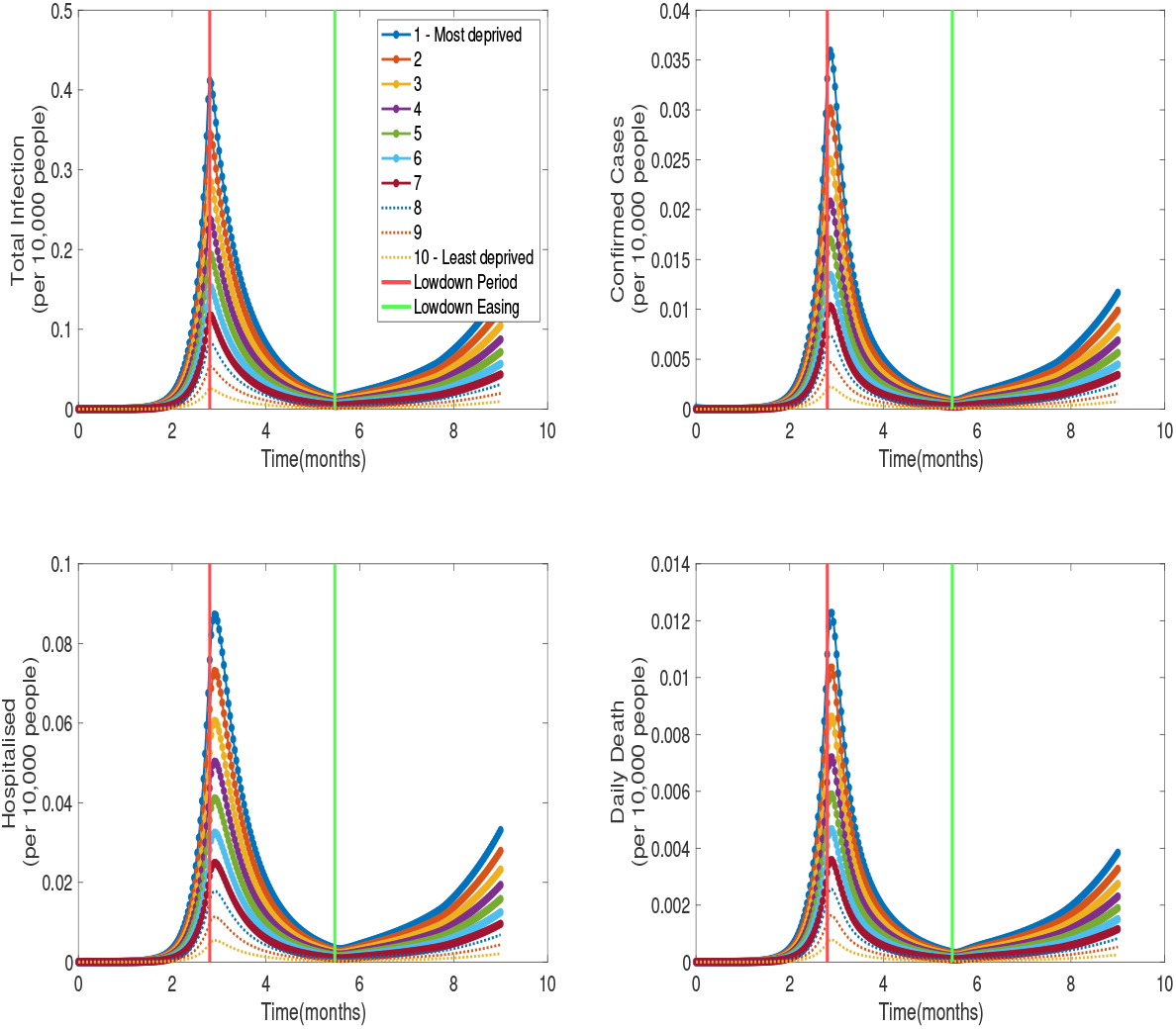
Time series of infections, reported cases, hospitalisations, and deaths per 10,000 individuals by deprivation decile, during the lockdown and easing period at *ϵ* = 0.7. **Top left panel** The daily number of infections by deprivation decile, **Top right panel** the number of reported cases, **Bottom right panel** the number of hospitalised individuals, **Bottom left panel** the number of daily deaths.

**Fig 5.**
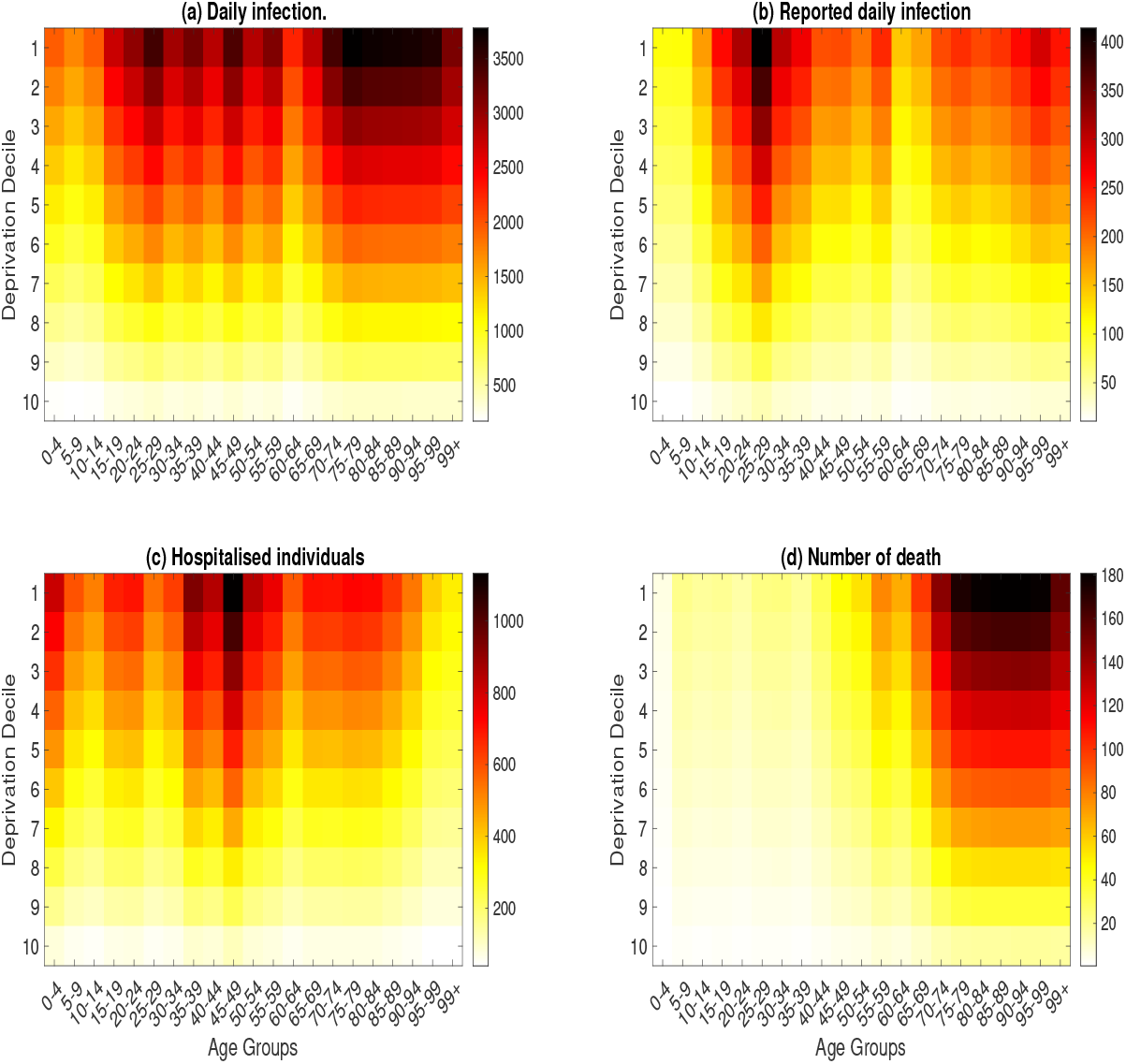
Aggregated daily infection, reported cases, hospitalizations, and deaths by age group and deprivation decile at *ϵ* = 1. **Top left panel:** sum of the daily number of infection aggregated by age and deprivation decile. **Top right panel:** sum of the daily number of reported cases aggregated by age and deprivation decile. **Bottom right panel:** The sum of hospitalised individuals by age and deprivation decile. **Bottom left panel** The sum of daily death by age and deprivation decile. This emphasizes the compounded effect of age and socioeconomic deprivation on COVID-19 outcomes, with the most deprived and oldest groups facing the highest risks of severe disease and death. This visualization underscores the need for targeted interventions to mitigate the disproportionate impact on vulnerable populations.

The proportionate mixing scenario would require a different type of control effort since the likelihood of interaction between social groups is the same. Hence, control effort would need to be targeted to the whole population and not a specific decile of deprivation. The diagonal mixing approach can give an inaccurate, underestimated, or overestimated view of the actual burden of the disease in each group, since although individuals usually interact within their social groups, there will still be some interactions with individuals from other groups that the diagonal mixing assumption did not take into account. Although both diagonal and proportion mixing models would require control effort targeted at the whole population, the proportionate assumption would require approximately 49% of the whole population compared to approximately 65% in diagonal mixing. This could be due to the fact that there is no interaction between groups in diagonal mixing, hence the requirement to target every group at a certain required threshold.

In contrast to prior research, our model enables the simultaneous inference of disease dynamics across social and age groups. By utilizing our model framework, it would be possible to examine the distinct age dynamics within each social group when data becomes accessible in the future, thus highlighting the impact of distinct age mixing patterns based on deprivation deciles. A constraint of this study is the lack of data required to estimate the average number of contacts within each deprivation decile. We suggest that forthcoming studies could adopt an approach similar to POLYMOD [25] to estimate the mixing structure among deprivation deciles in the UK. The main objective of this research is to showcase how mixing patterns impact the progression of the disease across deprivation deciles. Consequently, we postulate that the assumption of diagonal mixing is similar to homogeneous mixing within different social groups, yet it offers a straightforward method to collectively model these groups.

## Conclusion

In conclusion, this study aimed to investigate the influence of social mixing patterns on disease outcomes across heterogeneous populations. The results indicate a significantly higher rate of infection among the most deprived compared to the least deprived group of the population. Our model can facilitate the development of tailored non-pharmaceutical interventions targeting the most affected sociodemographic groups. This is achievable by using deprivation deciles as a proxy for sociodemographic groups, which indirectly enables spatial intervention due to the location-based nature of decile, as opposed to using income or ethnicity which can be more broadly representative of the entire population. We have developed a deterministic ordinary differential equation multi-risk structured model of COVID-19 disease outcomes, with a focus on the total number of infections, reported cases, hospitalised individuals and deaths in the population. We have provided a significant contribution by creating a mathematical model that considers age and sociodemographic categories, along with time-sensitive non-pharmaceutical interventions (such as lockdowns), to monitor the progression of a disease after control measures are put in place. Additionally, we incorporated a dynamic testing rate similar to logistic growth, which can be useful during a new outbreak of infection. This methodology could be applied in future studies to better understand the collective impact of age and socioeconomic status on the spread of the disease. This study has a limitation in the availability of data to parameterise the model and validate our assumptions. Despite the limitation, the model was designed to be more easily fitted to data when it becomes available. Additionally, our work will motivate future research to collect data that can be used to parameterise age-mixing dynamics in sociodemographic groups of interest. This would enable us to better understand the possible types of mixing that occur in each sociodemographic group by age.

## Data Availability

All data produced in the present study are available upon reasonable request to the authors

